# Differential pre-pandemic IgA reactivity against SARS-CoV-2 and circulating human coronaviruses measured in milk collected in Uganda and the USA

**DOI:** 10.1101/2021.06.24.21259294

**Authors:** Thomas G. Egwang, Tonny Jimmy Owalla, Emmanuel Okurut, Gonzaga Apungia, Alisa Fox, Claire DeCarlo, Rebecca L. Powell

**Affiliations:** Human Milk and Lactation Research Center, Med Biotech Laboratories, Kampala, Uganda; Department of Medicine, Division of Infectious Diseases, Icahn School of Medicine at Mount Sinai, New York, USA

## Abstract

**Objective:** Uganda, like other African countries, has registered fewer COVID-19 cases and deaths per capita than non-African countries. The lower numbers of cases and deaths in Uganda might be due to pre-existing cross-immunity induced by zoonotic coronaviruses or circulating common cold human coronaviruses (HCoVs) before the COVID-19 pandemic. In order to test this premise, we compared IgA reactivity to severe acute respiratory syndrome coronavirus 2 (SARS-CoV-2) and HCoVs in breast milk of US and rural Ugandan mothers collected in 2018 before the COVID-19 epidemic. Ugandan and US pre-pandemic breast milk samples were run in duplicate on enzyme-linked immunoadsorbent assay (ELISA) to measure specific IgA antibody reactivity to the spike proteins of SARS-CoV-2, human coronaviruses (HCoV) NL63, OC43, HKU1, and 229E. Pooled plasma from US COVID-19 positive and negative cases were employed as positive and negative controls, respectively. One Ugandan pre-pandemic milk sample had remarkably high reactivity against all HCoVs and SARS-CoV-2 spike proteins. There was higher IgA reactivity against the betacoronavirus HCoV-OC43 in Ugandan pre-pandemic milk samples by comparison with US pre-pandemic milk samples (p = 0.018). By contrast, there was significantly higher IgA reactivity against the alphacoronaviruses HCoV-229E and HCoV-NL63 in US pre-pandemic milk samples by comparison with Ugandan pre-pandemic milk samples (p < 0.0001 and 0.035, respectively).

**Conclusion:** Some Ugandan mothers may have robust pre-existing immunity against SARS-CoV-2 due to cross-immunity induced by HCoVs which may be passed on to their infants via breastfeeding. The differential pre-pandemic reactivity of US mothers to HCoV 229E and HCoV NL63 may have contributed to suboptimal antibody responses to SARS-CoV-2.

## 1. Introduction

Severe acute respiratory syndrome coronavirus 2 (SARS-CoV-2) has spread worldwide causing > 177,100,000 coronavirus disease 2019 (COVID-19) cases by June 18, 2021[1]. Uganda, like other African countries, has registered fewer COVID-19 cases and deaths per capita than non-African countries [1]. The lower numbers of cases and deaths in Africa by comparison with those in Western countries might be partly due to cross-immunity induced by circulating common cold human corona viruses (HCoVs) [2]. The HCoVs 229E, NL63, OC43 and HKU1 share high sequence similarity with SARS-CoV-2 proteins [3], cause mild upper respiratory tract infections (common cold) [4], and induce cross-reacting antibodies and T cells to SARS-CoV-2 in people never exposed to SARS-CoV-2 [5,6]. HCoV-induced antibodies with SARS-CoV-2 neutralizing activity targeting the S2 subunit of the SARS-CoV-2 Spike protein have been found to be present in pre-pandemic samples of 21/48 (44 %) children and 16/302 (5.3%) adults from the UK never exposed to SARS-CoV-2 [5]. A Dutch study similarly demonstrated pre-existing HCoV-specific T cells in young but not older adults [6]. Notably, the prevalence of HCoV-induced cross-reactive antibodies in pre-pandemic sub-Saharan African sera was 6-8-fold higher compared with US pre-pandemic sera [7]. These findings support the premise that systemic cross-immunity induced by HCoVs partly explains the lower numbers of COVID-19 cases and deaths in younger individuals in general and in Africa as a geographic region. However, there is a paucity of studies on pre-existing cross-reactive mucosal immunity against SARS-CoV-2.

Mucosal immunity is predominantly effected by IgA and secretory IgA (IgA/sIgA) and IgM/sIgM at mucosal surfaces such as the gastrointestinal and reproductive tracts, lungs, tears, saliva and breast milk [8]. There is increasing evidence of a robust mucosal immunity against SARS-CoV-2 [9]. IgA antibody responses to SARS-CoV-2 Spike protein have been reported in saliva [10]. Elevated IgA antibody responses against SARS-CoV-2 were reported in nasal fluids, tears, and saliva while elevated IgA-secreting plasmablasts expressing the mucosal chemokine receptor CCR10 were reported in peripheral blood of SARS-CoV-2-infected subjects [11,12]. CCR10 and its ligand CCL28 are uniquely involved in mucosal immunity [13,14]. Breast milk provides a unique window on mucosal immunity in mothers and provides protection against a legion of pathogens in infants [15]. There is evidence that breast milk IgA antibodies are robustly induced by natural SARS-CoV-2 infection and various COVID-19 vaccines. First, several studies demonstrated IgA antibodies against SARS-CoV-2 with virus neutralizing activity in breast milk of mothers with COVID-19 [16-18]. The reported breast milk antibody isotypes included IgA/secretory IgA(IgA/sIgA), IgM/sIgM, and IgG with IgA being the predominant antibody [16-19]. Second, COVID-19 vaccination has been rolled out in several countries and recent published reports indicate that vaccination induces robust IgA and IgG antibody responses with neutralizing activity in breast milk which may confer protection to breastfeeding infants [20-23].

The current limited access to COVID-19 vaccines in Africa leaves millions of mothers and their infants vulnerable to SARS-CoV-2 infection and COVID-19 related illnesses. Protection for these mother-infant pairs may therefore depend in the interim on pre-existing cross-reactive mucosal immunity acquired by mothers before the pandemic. There is a dearth of information about pre-existing cross-reactive mucosal immunity induced by HCoVs. Two US studies reported the presence of cross-reactive breast milk antibodies against HCoVs in US pre-pandemic breast milk samples which recognize and neutralize SARS-CoV-2 [16,17]. Pre-existing immunity in breastfeeding mothers might provide temporary protection of mother-infant dyads with no immediate access to COVID-19 vaccines. This is particularly relevant to sub-Saharan African countries where COVID-19 vaccination roll out is expected to be slow due to limited vaccine supplies and vaccine hesitancy must be overcome [25]. To the best of our knowledge, no study has investigated the presence of cross-reactive IgA antibodies against the S proteins of SARS-CoV-2 and HCoVs in pre-pandemic breast milk samples collected in Africa prior to 2019. In this study, we tested the hypothesis that breastfeeding rural Ugandan mothers who have never been exposed to SARS-CoV-2 have pre-existing cross-reactive IgA antibodies induced by endemic HCoVs which are passed to infants via breast milk. We therefore measured IgA antibodies against the S proteins of SARS-CoV-2 and the four HCoVs 229E, NL63, OC43 and HKU1 in pre-pandemic Ugandan milk samples collected in March 2018 before the emergence of the COVID-19 pandemic. We also compared IgA antibody responses in pre-pandemic Ugandan and US breast milk samples to gauge differences in pre-existing cross-reactive mucosal immunity between the two study populations before the pandemic. Our findings support the hypothesis that breastfeeding Ugandan mothers who have never been exposed to SARS-CoV-2 have pre-existing immunity which might be transferred to infants via breast milk.

## 2. Materials and Methods

### 2.1. Study samples

A cross-sectional study examining various breast milk parameters in a cohort of breastfeeding mother-infant dyads was undertaken in March 2018. The study was part of a larger malaria study that was reviewed and approved by the Research Ethics Committee of the Vector Control Division, Ministry of Health and the Uganda National Council for Science and Technology. The study was conducted at the Med Biotech Laboratories research clinic at St. Anne Health Center III in Usuk Subcounty, Katakwi District, in Northeastern Uganda. The study population comprised of mothers who were breastfeeding at the time of the study. Inclusion criteria were: (1) willingness to donate breast milk; (2) age 18 and above. The sole exclusion criterion was clinical suspicion or evidence of mastitis in one or both breasts. USA samples were collected in 2018-2019 in New York, NY from healthy women as part of studies related to influenza and HIV under approved Ichan School of Medicine at Mount Sinai IRB protocols. All donors provided written informed consent.

### 2.2. Milk collection

For Ugandan samples, breast milk was manually collected with the assistance of a lactation nurse as described [27]. Briefly, the skin around the areola and nipple was cleaned with an alcohol swab, the breast gently massaged, and 5-mL milk samples were manually expressed into 50-mL Falcon tubes which were immediately plunged into dry ice. Milk samples were transported to the lab in Kampala where they were aliquoted and stored at 20 °C until shipped on dry ice to the USA. For American samples, participants were asked to collect milk into a clean container using electronic or manual pumps at home. Milk was frozen in participants’ home freezers until samples were picked up by researchers and transferred on ice to the Mount Sinai Hospital where they were stored at -80°C until testing.

### 2.3. Enzyme-linked immunoadsorbent assay (ELISA)

To examine the levels of SARS-CoV-2 and HCoV Abs in human milk, we modified an ELISA that was recently developed and validated for use in blood serum/plasma [28,29] and have been successfully adapted this assay for use with human milk [18]. Briefly, before Ab testing, milk samples were thawed, centrifuged at 800g for 15 min at room temperature, fat was removed, and supernatant transferred to a new tube. Centrifugation was repeated 2x to ensure removal of all cells and fat. Skimmed acellular milk was aliquoted and frozen at -80°C until testing. Both Ugandan and US pre-pandemic milk samples were tested in duplicate, measuring IgA against the full trimeric Spike protein of SARS-CoV-2, or the S1 domain of HCoV-HKU1, HCoV-OC43, HCoV-229E, or HCoV-NL63. Due to the limited availability of Ugandan samples, milk was used diluted 1 in 10 and titrated 2.5-fold in 1% bovine serum albumin (BSA)/PBS and added to the plate. Plates were incubated at 4°C overnight, washed in 0.1% Tween 20/PBS (PBS-T), and blocked in PBS/3% goat serum/0.5% milk powder/3.5% PBS-T for 1h at room temperature. After 2h incubation at room temperature, plates were washed and incubated for 1h at room temperature with horseradish peroxidase-conjugated goat anti-human-IgA,(Rockland) diluted in 1% BSA/PBS. Plates were developed with 3,3’,5,5’-Tetramethylbenzidine (TMB) reagent followed by 2N hydrochloric acid (HCl) and read at 450nm on a BioTek Powerwave HT plate reader.

### 2.4. Data analysis

Endpoint dilution titers were determined from log-transformed titration curves using 4-parameter non-linear regression and an OD cutoff value of 1.0. Endpoint dilution positive cutoff values were determined as above. Mann-Whitney U tests were used to determine if the grouped Ugandan and American pre-pandemic milk samples differed in terms of specific reactivity to a given antigen. Pearson correlation tests were performed to compare SARS-CoV-2 and HCoV reactivity. All statistical tests were performed in GraphPad Prism, were 2-tailed, and significance level was set at p-values < 0.05.

## 3. Results

### 3.1. Pre-pandemic milk IgA reactivity to SARS-CoV-2

Pre-pandemic Ugandan and American milk samples (N=25 per group) were assayed in duplicate by ELISA in order to compare the levels of IgA antibody reactivity to the Spike protein. Pooled plasma from SARS-CoV-2 positive and negative donors were employed as positive and negative controls, respectively. Notably, 1 Ugandan sample exhibited very high reactivity to the SARS-CoV-2 Spike, as evident from the titration curve ODs and endpoint titer (Fig. 1A,F). In contrast, at the initial 1/10 dilution, grouped Ugandan and American samples exhibited similar reactivity (Ugandan mean OD=0.88, American mean OD=1.02, p=NS; Fig. 1A and data not shown). OD values at each milk dilution tested were used to determine endpoint titer. With the exception of the high-responder Ugandan sample, all other samples tested in both groups exhibited variable, low-level reactivity, with 16/25 samples in each group failing to reach the endpoint titer cutoff (OD=1.0; Fig. 1F). Ugandan samples exhibited a higher mean endpoint titer of 44.1 compared to that of American samples (12.7); however, this difference was not significant, as the outlying Ugandan high responder sample strongly influenced the higher mean (Fig. 1F).

**Figure 1.**
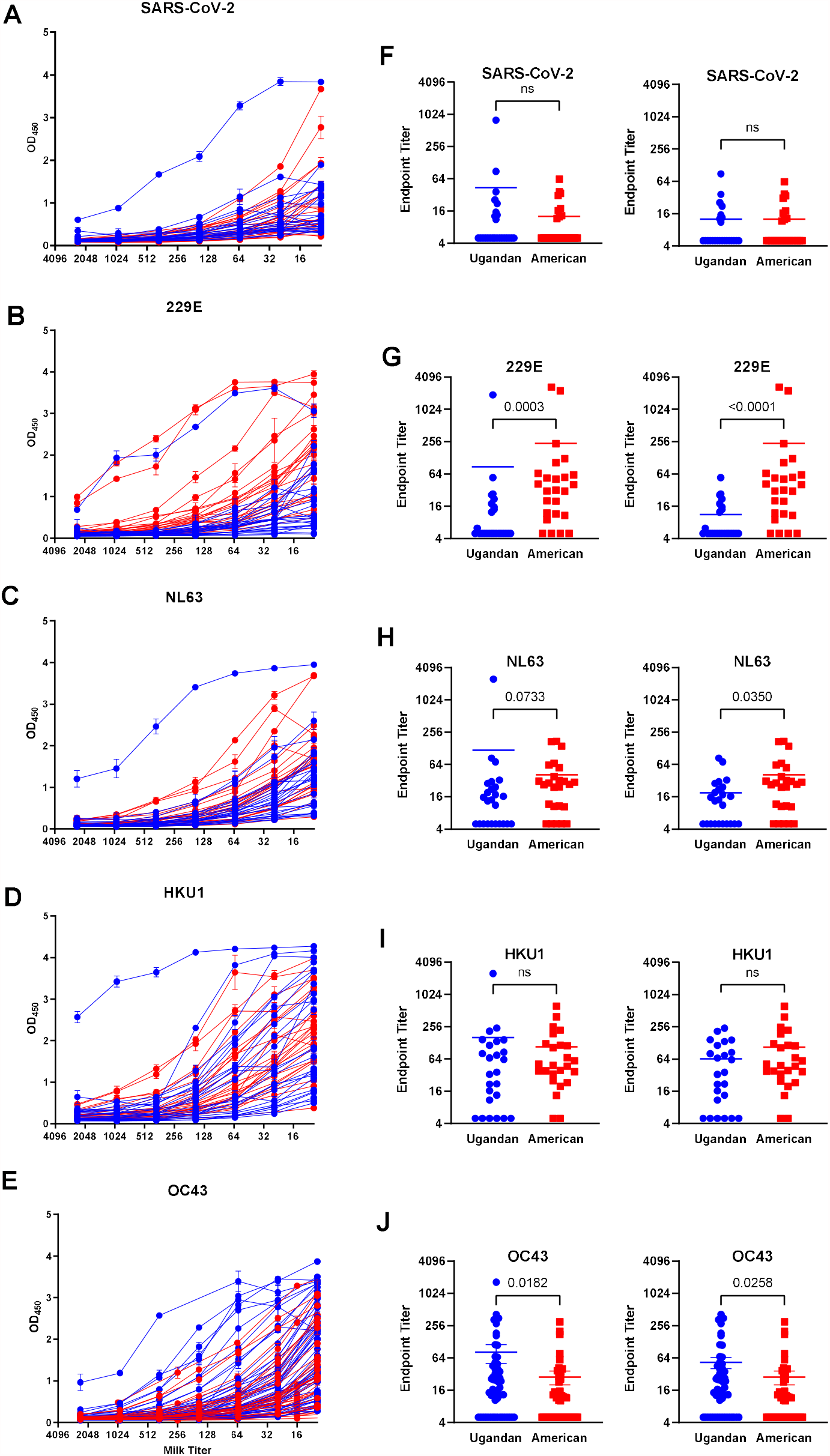
IgA Reactivity in pre-pandemic milk obtained from Ugandan and American donors against HCoVs and SARS-CoV-2. Milk was assayed by ELISA starting at 1/10 dilution. The full titrations against each Spike protein are shown in the first panel. Titration curves were used to determine endpoint titers using an OD cutoff of 1.0. Endpoint titers for each antigen tested are shown in the second panel. In the third panel, the Ugandan high responder was removed from the dataset, in order to compare differences without this outlier. Assays were performed in duplicate. Mean values of the scatter charts. Groups were compared by 2-tailed Mann-Whitney test; p values are shown.

### 3.2. Pre-pandemic milk IgA reactivity to HCoVs

#### 3.2.1. Human Alphacoronaviruses 229E and NL63

Milk samples were assayed against the HCoVs as described above. The Ugandan high-responder noted for SARS-CoV-2 was also highly reactive against both alphacoronaviruses (Fig. 1). In contrast, Ugandan milk samples at 1/10 dilution as a group exhibited significantly lower reactivity compared to American samples against HCoV-229E (Ugandan mean OD=0.87; American mean OD=1.95; p<0.0001) and HCoV-NL63 (Ugandan mean OD=1.27; American mean OD=1.62; p=0.0077; Fig. 1B,C and data not shown). Endpoint titers were significantly lower for the Ugandan samples compared to American samples against HCoV-229E (Ugandan mean endpoint titer=87.2; American mean endpoint titer=238; p=0.0003; Fig. 1G). Statistical significance of the difference was increased if the Ugandan high-responder was removed from the dataset (p<0.0001; Fig. 1G right panel). Against HCoV-NL63, endpoint titers were not significantly different if the Ugandan high responder was included in the data; however, if this outlier was removed, endpoint titers were also significantly lower for the Ugandan samples (Ugandan mean endpoint titer=18.2; American mean endpoint titer=41.4; p=0.035; Fig. 1H).

#### 3.2.2. Human Betacoronaviruses HKU1 and OC43

The Ugandan high-responder noted for SARS-CoV-2 was also highly reactive against both betacoronaviruses (Fig. 1). Against the HKU1 Spike, IgA reactivity was highly similar at 1/10 dilution between the Ugandan and American sample groups (Ugandan mean OD=2.16, American mean OD=2.26; p=NS; Fig. 1D and data not shown). Endpoint titers were also not significantly different, though the Ugandan mean was increased due to the robust response of the Ugandan high responder (Ugandan mean endpoint titer=162; American mean endpoint titer=107; p=NS). Against HCoV-OC43, initially it was evident that a subset of Ugandan samples exhibited increased reactivity, though comparing the 2 groups on the whole did not yield significant differences (data not shown); therefore, 30 additional Ugandan and 27 additional American samples were analyzed to determine if the observed trend of higher reactivity might become significant. It was found for milk at 1/10 dilution that Ugandan samples exhibited significantly higher IgA reactivity against HCoV-OC43 (Ugandan mean OD=1.76, American mean OD=1.37; p=0.0132; Fig. 1E and data not shown). Endpoint titers were also significantly higher for the Ugandan samples (Ugandan mean endpoint titer=82; American mean endpoint titer=28.3; p=0.0182; Fig. 1J). The higher reactivity of the Ugandan samples remained significant even with the removal of the high-responder from the dataset (p=0.0258; Fig.1J).

### 3.3. Correlation of HCoV and SARS-CoV-2 IgA reactivity

Correlation analyses were performed to determine the relationship between pre-pandemic HCoV and SARS-CoV-2 IgA reactivity for each group of milk samples. For the Ugandan milk samples, a significant positive correlation was found for reactivity against each HCoV compared to SARS-CoV-2 (Fig. 2A; p<0.0001). However, if the Ugandan high responder sample was removed from the dataset, these correlations were no longer evident (data not shown). No correlation between HCoV and SARS-CoV-2 IgA reactivity was found for the American samples (Fig. 2B).

**Figure 2.**
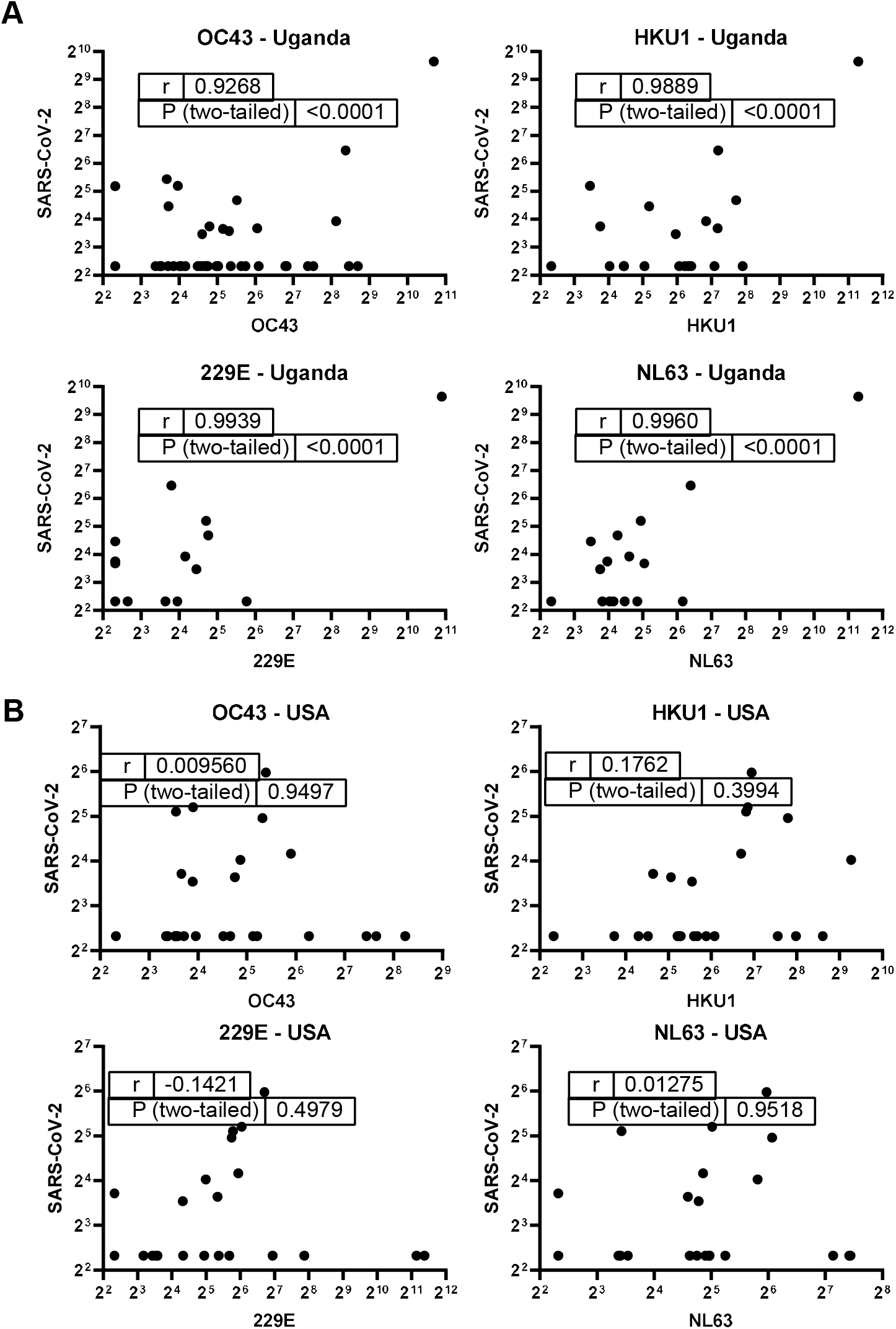
Correlation between HCoV and SARS-CoV-2 milk IgA reactivity. (A) Ugandan sample correlations. (B) American sample correlations. Endpoint titers were used for these analyses in 2-tailed Pearson correlation tests. P values are shown.

## 4. Discussion

Uganda has registered fewer COVID-19 cases and deaths per capita than the US and other Western countries [1]. The lower numbers of cases and deaths might be partly due to cross-immunity induced by circulating common cold HCoVs which share high sequence similarity with SARS-CoV-2 proteins [3] and induce cross-reacting antibodies and T cells to SARS-CoV-2 in people never exposed to SARS-CoV-2 [5,6]. Here we report for the first time that pre-pandemic breast milk samples obtained from rural Ugandan mothers in 2018 contain IgA antibodies that are reactive to the Spike proteins of all four HCoVs tested with low-to-moderate cross-reactivity to SARS-CoV-2. In comparing these data to those of pre-endemic American samples, it is evident that there is a considerable unique IgA reactivity profile for most of the HCoVs tested that is likely geographically-based.

Our analysis identified one Ugandan high-responder, and suggests that in addition to the common cold HCoVs, there might be zoonotic coronaviruses circulating in Uganda and other African countries that are highly related to SARS-CoV-2 that on rare occasions infect people but person-to-person transmission is either unlikely or the infections are asymptomatic and pass undetected. Indeed, molecular surveillance in Uganda and Kenya has identified bat coronaviruses which are relatives of the HCoVs Middle East respiratory syndrome coronavirus (MERS-CoV), NL63 and 229E [30,31]. A molecular investigation of a mild respiratory outbreak in wild chimpanzees in Cote d’Ívoire demonstrated human transmission of HCoV OC43 [32]. These studies underscore the potential bidirectional nature of HCoV transmission between wildlife and humans. The role of wildlife in high breast milk IgA cross-reactivity to HCoVs is not known. The Ugandan high responder is a donor who resides in a region in Northeastern Uganda known for livestock keeping. Livestock, wildlife, and humans share many pathogens [33]. However, milk samples donated by other women from the same region did not have high IgA cross-reactivity. It is possible that these women could have had higher milk antibody cross-reactivity in the other immunoglobulin isotypes which we did not investigate. It is also possible that zoonotic coronavirus infections are rare and affect only a tiny proportion of the population.

Molecular and serological surveys of HCoVs, MERS-CoV, and SARS-CoV-1 in humans has not been done in Uganda. A hospital-based study of Ugandan patients with influenza-like illness demonstrated a high seroprevalence of HCoVs but used a nucleoprotein-based ELISA which does not distinguish between different HCoVs [34]. A molecular surveillance in neighboring Kenya during 2009-2012 demonstrated overall HCoVs prevalence of 8.4 % among which HCoV OC43 predominated [35]. A molecular longitudinal study of seasonal coronaviruses in South African infants demonstrated an association between HCoV-OC43 and lower respiratory tract infections [36]. About one in two rural Ghanaians had IgG against three HCoVs with antibodies against HCoV-229E and HCoV-OC43 predominating [37]. A recent study targeting the nucleocapsid of SARS-CoV-2 and spike proteins of HCoVs reported a higher prevalence of antibodies against HCoVs in pre-pandemic African sera compared with US pre-pandemic sera, with cross-reactivity was attributed to the nucleocapsid protein of HCoV-NL63 [7]. As our study examined milk antibodies against only Spike proteins, our data cannot be readily compared to these serum studies; however, these previous studies and our own underscore the potential public health significance of certain HCoVs, especially OC43, in the context of pre-existing SARS-CoV-2 immunity.

Cross-reactive HCoV-OC43-specific mucosal immunity against SARS-CoV-2 could be providing moderate protection against COVID-19 in rural Uganda, despite the fact that many Ugandan samples did not exhibit increased milk IgA against SARS-CoV-2 compared to American samples. A significant correlation was present between IgA reactivity to SARS-CoV-2 and all 4 HCoVs when the Ugandan high responder was included, suggesting the presence of pre-existing immunity against SARS-CoV-2. Although the correlation did not hold true if this outlier was excluded, the existence of such outliers may be relevant on a population level in Uganda as there may be rare individuals with such potent cross-reactive immunity which impacts on the overall spread and severity of SARS-CoV-2. This premise is supported by several published studies. First, recent endemic HCoV infection is associated with a reduced severity of COVID-19 and risk of admission to the intensive care unit [38]. Second, previous viral symptoms correlated with the level and duration of cross-reactive antibodies to S1 and S2 subunits of SARS-CoV-2 and HCoV-OC43 in breast milk [39]. Third, higher antibody levels against HCoV-OC43 S protein were seen in individuals with pre-pandemic SARS-CoV-2 reactive antibodies than in those without [40]. Fourth, SARS-CoV-2 infection boosted antibodies reactive to the S2 domain of the HCoV-OC43 S protein [40]. Indeed, SARS-CoV-2 boosting of antibodies against other betacoronaviruses has been reported by others [41,42]. Additionally, HCoV-OC43 immunity may impact the cross-reactive response in ways not measured in the present study, including an enhanced memory B cell population and/or reactive T cells. It remains to be established how Ugandan milk IgA reactivity against HCoV-OC43 relates to mucosal immunity in other mucosal compartments such as the lungs which are most affected by severe COVID-19 disease. Remarkably, American pre-pandemic samples exhibited significantly more milk IgA reactivity to HCoV-229E and HCoV-NL63 compared to Ugandan samples. It is possible that this enhanced Alphacoronavirus response resulted in suboptimal cross-reactive SARS-CoV-2 antibodies. Suboptimal antibody responses can result in antibody dependent enhancement of infection and antibody-mediated immune enhancement with poor outcomes [43].

Our study has several limitations. First, only a small number of pre-pandemic samples were analyzed for IgA reactivity. It is therefore not possible to conclude whether a proportion of the Ugandan study population have similar cross-reactive immunity. Second, no SARS-CoV-2 neutralization studies were carried out to determine the functional relevance of the highly reactive IgA antibodies in some pre-pandemic Ugandan samples. We have previously reported that breast milk IgA titers correlate with virus neutralization activity in American mothers [44]. There are plans to identify Ugandan samples with high IgA reactivity and undertake neutralization experiments to establish their functional relevance to protection. Third, the cross-sectional design of the study precluded the generation of hypotheses about cause-effect relationships. The design was inevitable because samples were collected at a single time point before the COVID-19 pandemic. Fourth, the study only looked at IgA reactivity and did not investigate other antibody isotypes that may be relevant to mucosal cross-immunity against COVID-19. Finally, the study did not correlate breast milk and serum antibody reactivity to establish the relationship between systemic and mucosal cross-immunity to SARS-CoV-2 and HCoVs in the study region. Some of these limitations will be addressed in a future study with a larger number of Ugandan pre-pandemic breast milk samples.

## 5. Conclusion

In conclusion, our preliminary findings support the premise that mucosal HCoV immunity varies geographically, and a proportion of rural Ugandan mothers who have not been exposed to SARS-CoV-2 may have pre-existing cross-immunity induced by certain HCoVs or zoonotic coronaviruses. This cross-immunity might protect rural mothers and their infants against COVID-19. These results have public health significance and are generalizable to rural mothers in other African countries. COVID-19 vaccination rollout is expected to be slow in Africa and therefore pre-existing immunity may help prevent or reduce the severity of COVID-19 in unvaccinated populations during the interim.

## Data Availability

All data reffered to in the manuscript are available on reasonable request

## Authorship contribution statement

Thomas Egwang: Funding acquisition, Resources, Supervision, Writing-review & editing. Tonny Owalla: Methodology. Emmanuel Okurut: Methodology. Gonzaga Apungia: Methodology. Alisa Fox: Methodology. Claire DeCarlo: Methodology. Rebecca Powell: Funding acquisition, Investigation, Supervision. Methodology, Formal analysis, Writing-review & editing.

## Acknowledgments

We thank the mothers of Abwokodia Parish, Osuk subcounty, and NYC who donated breast milk. Funding: This work was supported by Grand Challenges Canada and Global Innovation Fund grant 0817-05 (to T.G.E.) and by the Icahn School of Medicine at Mount Sinai (to R.P.). The funders played no role in the study design, data collection and analysis, and manuscript preparation.

## References

[1] WHO Coronavirus Disease (COVID-19) Dashboard. Accessed on June 18, 2021. https://covid19.who.int/?gclid=EAIaIQobChMI6OvIh-D06QIVVYfVCh1y9AotEAAYASAAEgJemfD_BwE.

[2] P. Doshi, Covid-19: Do many people have pre-existing immunity? BMJ. 370 (2020) m3563, https://doi:10.1136/bmj.m3563.

[3] N. Kaur, R. Singh, Z. Dar, R.K. Bijarnia, N. Dhingra, T. Kaur, Genetic comparison among various coronavirus strains for the identification of potential vaccine targets of SARS-CoV2. Infect Genet Evol. (2020) 104490, https://doi:10.1016/j.meegid.2020.104490.

[4] L. van der Hoek, Human coronaviruses: what do they cause? Antivir Ther. 12 (2007) 651–8.

[5] K.W. Ng, N. Faulkner, G.H. Cornish, A. Rosa, R. Harvey, S. Hussain, et al., Preexisting and de novo humoral immunity to SARS-CoV-2 in humans. Science. 370 (2020) 1339–1343, https://doi:10.1126/science.abe1107.

[6] G. Saletti, T. Gerlach, J.M. Jansen, A. Molle, H. Elbahesh, M. Ludlow, et al, Older adults lack SARS CoV-2 cross-reactive T lymphocytes directed to human coronaviruses OC43 and NL63. Sci Rep. 10(1) (2020) 21447. doi: 10.1038/s41598-020-78506-9. PMID: 33293664; PMCID: PMC7722724.

[7] F.Y. Tso, S.J. Lidenge, P.B. Peña, A.A. Clegg, J.R. Ngowi, J. Mwaiselage, et.al., High prevalence of pre-existing serological cross-reactivity against severe acute respiratory syndrome coronavirus-2 (SARS-CoV-2) in sub-Saharan Africa. Int J Infect Dis. 102 (2021) 577–583, https://doi:10.1016/j.ijid.2020.10.104.

[8] P. Brandtzaeg, The mucosal immune system and its integration with the mammary glands. J Pediatr. 156 (2 Suppl) (2010) S8–15, https://doi:10.1016/j.jpeds.2009.11.014.

[9] M.W. Russell, Z. Moldoveanu, P.L. Ogra, J. Mestecky, Mucosal immunity in COVID-19: a neglected but critical aspect of SARS-CoV-2 infection. Front Immunol. 11(2020) 611337, https://doi:10.3389/fimmu.2020.611337.

[10] Isho B, Abe KT, Zuo M, Jamal AJ, Rathod B, Wang JH, et.al., Persistence of serum and saliva antibody responses to SARS-CoV-2 spike antigens in COVID-19 patients. Sci Immunol. 5(2020) eabe5511, https://doi:10.1126/sciimmunol.abe5511.

[11] C. Cervia, J. Nilsson, Y. Zurbuchen, A. Valaperti, J. Schreiner, A. Wolfensberger, et.al., Systemic and mucosal antibody responses specific to SARS-CoV-2 during mild versus severe COVID-19. J Allergy Clin Immunol. 147(2021) 545–557.e9, https://doi:10.1016/j.jaci.2020.10.040.

[12] D. Sterlin, A. Mathian, M. Miyara, A. Mohr, F. Anna, L. Claër, et.al., IgA dominates the early neutralizing antibody response to SARS-CoV-2. Sci Transl Med. 13(2021) eabd2223, https://doi:10.1126/scitranslmed.abd2223.

[13] N. Xiong, Y. Fu, S. Hu, M. Xia, J. Yang, CCR10 and its ligands in regulation of epithelial immunity and diseases. Protein Cell. 3(8) (2012) 571–80.

[14] O. Morteau, C. Gerard, B. Lu, S. Ghiran, M. Rits, Y. Fujiwara, et.al., An indispensable role for the chemokine receptor CCR10 in IgA antibody-secreting cell accumulation. J Immunol. 181(9) (2008) 6309–15.

[15] K. Le Doare, B. Holder, A. Bassett, P.S. Pannaraj, Mother’s Milk: A Purposeful Contribution to the Development of the Infant Microbiota and Immunity. Front Immunol. 9 (2018) 361, https://doi:10.3389/fimmu.2018.00361.

[16] R.M. Pace, J.E. Williams, K.M. Järvinen, M.B. Belfort, C.D.W. Pace, K.A. Lackey, et.al., Characterization of SARS-CoV-2 RNA, Antibodies, and Neutralizing Capacity in Milk Produced by Women with COVID-19. mBio. 12 (2021) e03192–20, https://doi:10.1128/mBio.03192-20.

[17] V. Demers-Mathieu, D.M. Do, G.B. Mathijssen, D.A. Sela, A. Seppo, K.M. Järvinen, E. Medo, Difference in levels of SARS-CoV-2 S1 and S2 subunits-and nucleocapsid protein-reactive SIgM/IgM, IgG and SIgA/IgA antibodies in human milk. J Perinatol. 41(2021) 850–859, https://doi:10.1038/s41372-020-00805-w.

[18] A. Fox, J. Marino, F. Amanat, F. Krammer, J. Hahn-Holbrook, S. Zolla-Pazner, R.L. Powell, Robust and Specific Secretory IgA Against SARS-CoV-2 Detected in Human Milk. iScience. 23 (2020) 101735, https://doi:10.1016/j.isci.2020.101735.

[19] F. Zhu, C. Zozaya, Q. Zhou, C. De Castro, P.S. Shah, SARS-CoV-2 genome and antibodies in breastmilk: a systematic review and meta-analysis. Arch Dis Child Fetal Neonatal Ed. (2021) F1–F8, https://doi:10.1136/archdischild-2020-321074.

[20] A.Y. Collier, K. McMahan, J. Yu, L.H. Tostanoski, R. Aguayo, J. Ansel, et.al., Immunogenicity of COVID-19 mRNA Vaccines in Pregnant and Lactating Women. JAMA. (2021), https://doi:10.1001/jama.2021.7563.

[21] S.H. Perl, A. Uzan-Yulzari, H. Klainer, L. Asiskovich, M. Youngster, E. Rinott, I. Youngster, SARS-CoV-2-Specific Antibodies in Breast Milk After COVID-19 Vaccination of Breastfeeding Women. JAMA. (2021) e215782, https://doi:10.1001/jama.2021.5782.

[22] N.O. Shlomai, Y. Kasirer, T. Strauss, T. Smolkin, R. Marom, E.S. Shinwell, et.al., Neonatal SARS-CoV-2 Infections in Breastfeeding Mothers. Pediatrics. 147 (2021) e2020010918, https://doi:10.1542/peds.2020-010918.

[23] K.J. Gray, E.A. Bordt, C. Atyeo, E. Deriso, B. Akinwunmi, N. Young, et.al., COVID-19 vaccine response in pregnant and lactating women: a cohort study. Am J Obstet Gynecol. S0002-9378(2021)00187-3, https://doi:10.1016/j.ajog.2021.03.023.

[24] K.A. Callow, Effect of specific humoral immunity and some non-specific factors on resistance of volunteers to respiratory coronavirus infection. J Hyg (Lond). 95 (1985)173–89, https://doi:10.1017/s0022172400062410.

[25] F.S. Wirsiy, C.N. Nkfusai, D.E. Ako-Arrey, E.K. Dongmo, F.T. Manjong, S.N. Cumber, Acceptability of COVID-19 Vaccine in Africa. Int J MCH AIDS. 10 (2021)134–138, https://doi:10.21106/ijma.482. Epub 2021 Apr 8. PMID: 33868778; PMCID: PMC8039868.

[26] T.J. Owalla, E. Okurut, G. Apungia, B. Ojakol, J. Lema, S.C. Murphy, T. Egwang, Using the ultrasensitive Alere Plasmodium falciparum Malaria Ag HRP-2<sup>™</sup> Rapid Diagnostic Test in the field and clinic in Northeastern Uganda. Am J Trop Med Hyg. 103 (2020)778–784, https://doi:10.4269/ajtmh.19-0653.

[27] T.M. Slusher, I.L. Slusher, E.M. Keating, B.A. Curtis, E.A. Smith, E. Orodriyo, et.al., Comparison of maternal milk (breastmilk) expression methods in an African nursery. Breastfeed Med. 7(2012) 107–11, https://doi:10.1089/bfm.2011.0008.

[28] F. Amanat, D. Stadlbauer, S. Strohmeier, T.H.O. Nguyen, V. Chromikova, M. McMahon, et.al., A serological assay to detect SARS-CoV-2 seroconversion in humans. Nat Med. 26 (2020) 1033–1036.

[29] D. Stadlbauer, F. Amanat, V. Chromikova, K. Jiang, S. Strohmeier, G.A. Arunkumar, et.al., SARS-CoV-2 Seroconversion in Humans: A Detailed Protocol for a Serological Assay, Antigen Production, and Test Setup. Curr Protoc Microbiol. 57(2020) e100, https://doi:10.1002/cpmc.100.

[30] S.J. Anthony, K. Gilardi, V.D. Menachery, T. Goldstein, B. Ssebide, R. Mbabazi R, et al., Further Evidence for Bats as the Evolutionary Source of Middle East Respiratory Syndrome Coronavirus. mBio. 8 (2017) e00373–17, https://doi:10.1128/mBio.00373-17.

[31] Y. Tao, M. Shi, C. Chommanard, K. Queen, J. Zhang, W. Markotter, et al, Surveillance of Bat Coronaviruses in Kenya Identifies Relatives of Human Coronaviruses NL63 and 229E and Their Recombination History. J Virol. 91(2017) e01953–16, https://doi:10.1128/JVI.01953-16.

[32] L.V. Patrono, L. Samuni, V.M. Corman, L. Nourifar, C. Röthemeier, R.M. Wittig, et.al., Human coronavirus OC43 outbreak in wild chimpanzees, Côte d’Ivoire, 2016. Emerg Microbes Infect. 7 (2018)118, https:/doi:10.1038/s41426-018-0121-2.

[33] J.L. Siembieda, R.A. Kock, T.A. McCracken, S.H. Newman, The role of wildlife in transboundary animal diseases. Anim Health Res Rev. 12 (2011) 95–111, https://doi:10.1017/S1466252311000041. Epub 2011 May 27. PMID: 21615975.

[34] E. Mulabbi, R. Tweyongyere, F. Wabwire, E. Mworozi, J. Koehlerb, H. Kibuuka, et al., Seroprevalence of human coronaviruses among patients visiting Hospital-Based Sentinel Sites in Uganda. Preprint (2020), https:/doi:10.21203/rs.3.rs-116084/v1.

[35] L.A. Sipulwa, J.R. Ongus, R.L. Coldren, W.D. Bulimo, Molecular characterization of human coronaviruses and their circulation dynamics in Kenya, 2009-2012. Virol J. 13 (2016)18, https://doi:10.1186/s12985-016-0474-x.

[36] M.P. Nicol, R. MacGinty, L. Workman, J.A.M. Stadler, L. Myer, V. Allen, et al., A Longitudinal Study of the Epidemiology of Seasonal Coronaviruses in an African Birth Cohort. J Pediatric Infect Dis Soc. (2021) piaa168, https://doi:10.1093/jpids/piaa168.

[37] M. Owusu, A.A. Sylverken, P. El-Duah, G. Acheampong, M.Y. Mutocheluh, Y. Adu-Sarkodie, Sero-epidemiology of human coronaviruses in three rural communities in Ghana. Pan African Medical Journal. 2021;38:244. [doi: 10.11604/pamj.2021.38.244.26110].

[38] Sagar M, Reifler K, Rossi M, Miller NS, Sinha P, White LF, et.al., Recent endemic coronavirus infection is associated with less-severe COVID-19. J Clin Invest. 131(2021) 131(1):e143380, https://doi:10.1172/JCI143380.

[39] Demers-Mathieu V, DaPra C, Mathijssen GB, Medo E. Previous viral symptoms and individual mothers influenced the leveled duration of human milk antibodies cross-reactive to S1 and S2 subunits from SARS-CoV-2, HCoV-229E, and HCoV-OC43. J Perinatol. 2021 May;41(5):952–960. doi:10.1038/s41372-021-01001-0. Epub 2021 Mar 1.

[40] E.M. Anderson, E.C. Goodwin, A. Verma, C.P. Arevalo, M.J. Bolton, M.E. Weirick, et.al., Seasonal human coronavirus antibodies are boosted upon SARS-CoV-2 infection but not associated with protection. Cell. 184 (2021) 1858–1864.e10, https://doi:10.1016/j.cell.2021.02.010.

[41] P. Nguyen-Contant, A.K. Embong, P. Kanagaiah, F.A. Chaves, H. Yang, A.R. Branche, et al., S Protein-Reactive IgG and Memory B Cell Production after Human SARS-CoV-2 Infection Includes Broad Reactivity to the S2 Subunit. mBio. 11 (2020)11 e01991-20, https://doi:10.1128/mBio.01991-20.

[42] E. Shrock, E. Fujimura, T. Kula, R.T. Timms, I.H. Lee, Y. Leng, et al., Viral epitope profiling of COVID-19 patients reveals cross-reactivity and correlates of severity. Science 370(2020) eabd4250, https://doi:10.1126/science.abd4250.

[43] A. Iwasaki, Y. Yang, The potential danger of suboptimal antibody responses in COVID-19. Nat Rev Immunol. 20 (2020) 339–341, https://doi:10.1038/s41577-020-0321-6.

[44] A. Fox, J. Marino, F. Amanat, K. Oguntuyo, J. Hahn-Holbrook J, B. Lee, et al,, The spike-specific IgA in milk commonly-elicited after SARS-Cov-2 infection is concurrent with a robust secretory antibody response, exhibits neutralization potency strongly correlated with IgA binding, and is highly durable over time. medRxiv preprint (2021) https://doi.org/10.1101/2021.03.16.21253731.

